# A prospective longitudinal study of chronic pulmonary aspergillosis in pulmonary tuberculosis in Indonesia (APICAL)

**DOI:** 10.1101/2021.04.25.21256062

**Authors:** Findra Setianingrum, Anna Rozaliyani, Robiatul Adawiyah, Ridhawati Syam, Mulyati Tugiran, Cut Yulia I. Sari, Finny Nandipinto, Johannes Ramnath, Arief Riadi Arifin, Diah Handayani, Erlina Burhan, Martin C. Rumende, Retno Wahyuningsih, Riina Rautemaa-Richardson, David W. Denning

## Abstract

**Objectives:** Chronic pulmonary aspergillosis (CPA) can complicate recovery from pulmonary tuberculosis (TB). CPA may also be misdiagnosed as bacteriologically-negative TB. This study aimed to determine the incidence of CPA in patients treated for TB in Indonesia; a country with a high incidence of TB.

**Methods:** In this prospective, longitudinal cohort study in patients treated for pulmonary TB, clinical, radiological and laboratory findings were analysed. Sputum was collected for fungal culture and TB PCR. Patients were assessed at baseline (0-8 weeks) and at the end (5-6 months) of TB therapy. CPA diagnosis was based on symptoms (≥3 months), characteristic radiological features and positive *Aspergillus* serology, and categorized as proven, probable and possible.

**Results:** Of the 216 patients recruited, 128 (59%) were followed up until end of TB therapy. At baseline, 91 (42%) had microbiological evidence for TB. *Aspergillus*-specific IgG was positive in 64 (30%) patients and went from negative to positive in 16 (13%) patients during TB therapy. The incidence of proven and probable CPA at baseline was 6% (n=12) and 2% (n=5) and end of TB therapy 8% (n=10) and 5% (n=7), respectively. Six patients (2 with confirmed TB) developed an aspergilloma. Diabetes mellitus was a significant risk factor for CPA (p=0.040). Persistent cough (n=5, 50%; p=0.005) and fatigue (n=6, 60%; p=0.001) were the most common symptoms in CPA.

**Conclusion:** CPA should be considered a relatively frequent differential diagnosis in patients with possible or proven TB in Indonesia. Lack of awareness and limited access to *Aspergillus*-specific IgG tests and CT imaging are obstacles in establishing a CPA diagnosis.

**Key messages:** *What is the key question?:* Do what extent is chronic pulmonary aspergillosis (CPA) both a) mistaken for TB and b) co-exists with TB during the course of 6 months therapy

*What is the bottom line?:* Features consistent with CPA were present in 6% of patients when starting TB therapy and 8% at the end of therapy, with some resolving and some developing CPA de novo during TB therapy. At the end of B therapy symptoms, cavitations with *Aspergillus*-specific IgG detectable were the key features of CPA.

*Why read on?:* Co-existence of TB and CPA is present in a substantial minority of patients starting and ending TB therapy, and needs addressing in terms of diagnosis, dual therapy and follow up.

## Introduction

The link between pulmonary TB and the subsequent development CPA is well established for those who have recovered from TB [1,2]. Studies to date have addressed the relative rate of development of CPA with slightly different incidence rates depending on the modes of testing, timing, diagnostic criteria used and the patient population [3–5]. No prospective studies have yet addressed the occurrence of both CPA and TB during anti-tuberculous (TB) therapy, although a few such cases are described [6]. Mis-diagnosis of CPA as pulmonary TB is relatively common in clinical practice, but hard to quantify as it is difficult to absolutely rule TB out as a diagnosis [5]. No prospective studies of CPA have been done in Indonesia. In 2019, Indonesia reported 845,000 TB cases to the WHO, 87% pulmonary [7] so the numerical impact of mis-diagnosis could be large.

Once a diagnosis of pulmonary tuberculosis is considered based on clinical symptoms, disease duration and radiological appearance, the standard approach and general clinical recommendation is to commence anti-TB therapy. In a review of older studies before the HIV era and before effective therapy for pulmonary TB was available [8], Tiemersma found that even in those with smear positive TB, the spontaneous recovery was 30% (14-47%) and in those with smear negative TB was ∼80%. So the drive to immediately treat patients with suspected TB is driven by concern for transmissibility, rather than immediate death of the patient. In the era of co-infection of HIV and TB, the 1 year mortality is still excessively high, but this is not the predominant problem in Indonesia [9].

This longitudinal study was designed to address the timing of the development of CPA during the course of treated tuberculosis. A very common feature of TB is pulmonary cavitation, and we know that this is an important precursor to CPA [2,10], and given that a) *Aspergillus* inhalation is inevitable and daily and b) that some genetic variants of humans are strongly associated with CPA, it would seem likely that some patients might develop CPA early in the course of TB [11,12]. This early *Aspergillus* infection could be progressive, or it could be self-limited in those with adequate immune defenses, and in both cases be marked by a raised *Aspergillus* antibody. This is not currently known [13–15].

Furthermore, in those with a clinical presentation of TB, it may be that some patients do not have TB, but an alternative diagnosis. The differential diagnosis includes CPA, nontuberculous mycobacterial infection, chronic cavitary pulmonary histoplasmosis, coccidioidomycosis or paracoccidioidomycosis, pulmonary cryptococcal infection, necrotizing lung cancer, pulmonary hydatid disease, actinomycosis, or a sub-acute bacterial infection on a background of bullous emphysema which appears radiologically similar to TB [16–19]. Histoplasmosis is endemic in Indonesia, but coccidioidomycosis and paracoccidioidomycosis are not, so only international travel would allow these infections to be manifest [20]. We hypothesized that some smear and TB PCR negative patients might have CPA and not TB, as described in Nigeria by Oladele [5]. By excluding patients with those previously treated for TB (based on the medical records and verbal confirmation from patients), we have taken out of consideration other populations of patients who might have CPA or a broader differential diagnosis.

Our primary objective was to determine the incidence of CPA during the course of and at the end TB therapy, and to assess if the criteria for CPA diagnosis are suitable for this population of patients who are already ill with TB. A second objective was to assess how many patients who are thought to have TB, but lack microbiological evidence for this, actually have CPA and how many have dual infections. We also hoped to determine if any patients had ‘self-resolving CPA’, or ‘sub-clinical CPA’ which does not progress, entities not yet adequately described in the literature, and which challenge the current general concept of CPA as a progressive disorder [21]. Our hypotheses are that: CPA is common and under-diagnosed in TB patients in Indonesia, there is dual infection between CPA and TB, CPA starts to develop during active infection of TB, and the current diagnostic criteria for resource-constrained settings are applicable in Indonesia.

## Methods

### Study population

This was a prospective longitudinal study Patients were enrolled at two tertiary care hospitals (National Referral Centre Persahabatan Hospital and National Referral Centre Cipto Mangunkusumo Hospital), and 4 district hospitals (Grha Permata Ibu Hospital, Universitas Kristen Indonesia Hospital, Cempaka Putih Jakarta Islamic Hospital, and MH Thamrin Hospital), serving greater Jakarta (12 million). Patients were referred from primary care because of diagnostic uncertainty, patient request or co-morbidity.

All consecutive consenting newly diagnosed pulmonary TB patients were recruited into the study. The exclusion criteria were previous history of TB, multi drug-resistant TB, less than 16 years of age, and non-consenting patients. Ethical approval was obtained from the University Research Ethics Committee, University of Manchester (approval number 16416) and from the ethics committee of the Faculty of Medicine, Universitas Indonesia (approval number 672/UN2.F1/ETIK/2016).

Pulmonary tuberculosis was diagnosed by the attending clinical team based on typical symptoms, positive TB PCR by GeneXpert MTB/RIF (Cepheid, CA, USA) and/or acid fast bacilli (AFB) smear and/or radiology finding positive for TB, as per national guidelines [22]. Culture for *Mycobacterium* spp. was not done, except in exceptional circumstances. Many patients were treated empirically if the suspicion for pulmonary TB was high.

### Data collection

Patients were assessed at the start of TB therapy (baseline, 0-8 weeks) and at the end of TB therapy (at 5-6 months). The main variables, collected at both time points, were serum *Aspergillus-*specific IgG levels, symptoms and quality of life scores.

A validated quality of life instrument used in respiratory disease – the St Georges Respiratory Questionnaire (SGRQ) was used in addition to CPA-specific questions [23–25]. This questionnaire consists of 3 domains (activity, impact and symptoms, each with range of 1-100) and high scores reflect worse quality of life.

### Sample collection and processing

Blood samples (5-10ml) and sputum (if produced) was collected from patients in the clinic at both time points. Aspergillus-specific IgG level was measured using the Immulite 2000 system (Siemens, Berlin, Germany) according to the manufacturer’s instructions using a cutoff of 11.5 mg/l [26]. Prior to testing, serum was stored in -80°C. Sputum from patients with productive cough was tested for TB using GeneXpert MTB/RIF PCR (Cepheid, CA, USA). Acid fast bacilli (AFB) smear results were collected from patients’ medical records. Any remaining sputum was cultured for fungi.

### Radiological interpretation

All patients had a chest radiograph at baseline and end of TB therapy. CT scans were also done for patients who could afford them. All images were digitized and reviewed by an experienced radiologist (FN) who coded the appearances systematically and provided a narrative report. All radiographs were also reviewed one of the authors (DWD) who specializes in aspergillosis. All radiographs were interpreted in the absence of data on symptoms or any laboratory findings or the interpretation of the other reviewer. The presence of pleural thickening adjacent to a cavity was required to suspect CPA, as opposed to anywhere on the radiograph -the only finding where there was some difference between the two reviewers who were in agreement for all other criteria.

### Definitions

Proven CPA was diagnosed based on three criteria: 1) at least one of the following symptoms; cough, haemoptysis, chest pain, dyspnea, fatigue, and/or weight loss ≥3 months, AND 2) radiological features indicative of CPA (cavitation, fungal ball or pleural thickening) and/or progressive cavitation on serial chest radiographs AND 3) positive *Aspergillus* IgG and/or *Aspergillus* spp detected by culture. The diagnosis of probable CPA was made when criteria 1 and 2 but not 3 were met AND 4) the PCR for *M. tuberculosis* was negative. Possible CPA was diagnosed in those who met criteria 1 and 2 but not 3 or 4. Criteria for proven, probable and possible CPA were modified versions from Denning et al [15]. “CPA” in the tables and text refers to proven CPA, unless otherwise stated.

### Data analysis

Statistical analysis was performed with the use of IBM SPSS 25 statistic software. A p value <0.05 was considered statistically significant. Data were presented using frequencies and percentages for binary and categorical variables, medians and range for non-normally distributed, continuous variables. The difference between non-parametric continuous variables was analysed using Mann-Whitney U-test for CPA and non CPA groups. Fisher’s exact tests or chi-squared tests were used for categorical variables for CPA and non CPA groups. Comparisons of median results across different time points were assessed by Wilcoxon signed-rank test. In addition, post hoc analyses were carried out with McNemar’s test with adjusted Bonferonni corrections between a two time-point comparisons.

## Results

A total of 216 patients with pulmonary TB were recruited at the start of their TB therapy and 128 (59%) patients re-attended the end of TB therapy follow-up appointment (Figure 1). The recruitment phase for the baseline appointments is from February 2017 until April 2018. At baseline, 91 (42%) of 216 patients had microbiological evidence for TB (positive TB PCR and/or AFB smear), 91 (42%) patients were clinically diagnosed as TB but had no microbiological evidence of it (negative TB PCR and/or AFB), and 34 (16%) patients were clinically diagnosed as TB but microbiological diagnostics had not been performed. An additional 2 patients had a positive TB PCR result at the end of TB therapy. Twelve (6%) patients met the criteria of proven CPA, five (2%) patients as probable CPA, and 15 (7%) patients as possible CPA, an incidence of 7.9% (95%CI: 4.7, 12.3) or 14.4% if possible cases are included. Six patients grew *Aspergillus* in sputum culture, two of whom met the criteria of possible CPA. None of the patients with proven CPA grew *Aspergillus* in their sputum.

**Figure 1.**
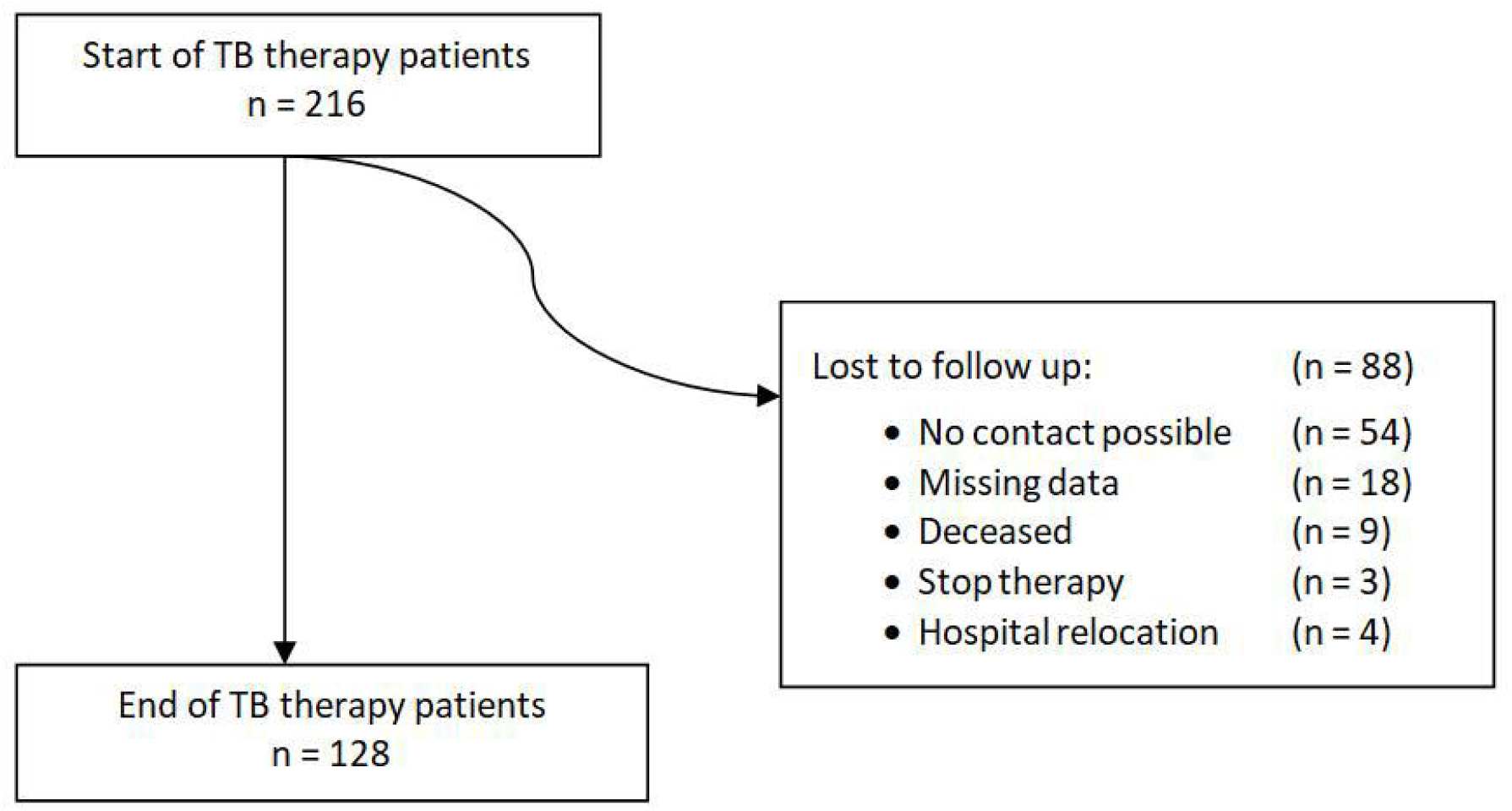
Overview of the study population

Males outnumbered females in CPA cases with a 3:1 ratio (Table 1). Diabetes mellitus appeared to be a significant risk factor for CPA at baseline assessment (p = 0.040, RR = 3.346, 42% in CPA patients vs 16% in non-CPA patients). There was significant difference in the proportion of cough (93% in CPA patients vs 13% in non-CPA patients), fatigue (42% in CPA patients vs 8% in non-CPA patients), and weight-loss (17% in CPA patients vs 1% in non-CPA patients) between CPA and non-CPA cases (p < 0.05). Baseline quality of life SGRQ scores are shown in Table 2. There was a significant difference between CPA and non-CPA patients in SQRQ in the symptoms domain [p=0.029, median 58 (CPA) vs 39 (non-CPA)]. Infiltrates (58% in CPA patients vs 87% in non-CPA patients), cavitation (92% in CPA patients vs 52% in non-CPA patients), air fluid level in cavities (50% in CPA patients vs 22% in non-CPA patients) and pleural thickening (84% in CPA patients vs 27% in non-CPA patients); all were more common in CPA cases (p <0.05) (Table 1).

**Table 1.**
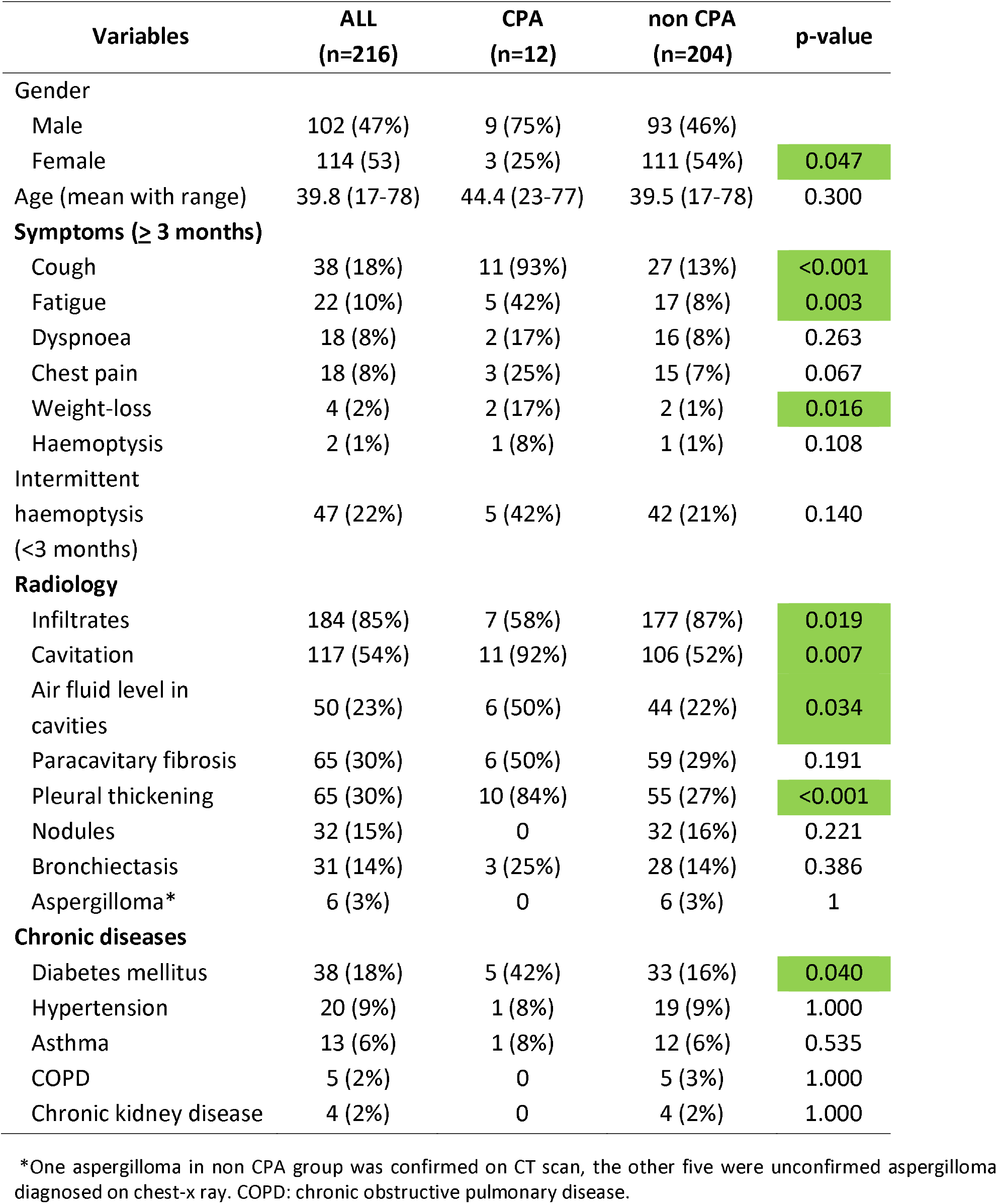
Patients’ characteristics at baseline assessment.

**Table 2.**
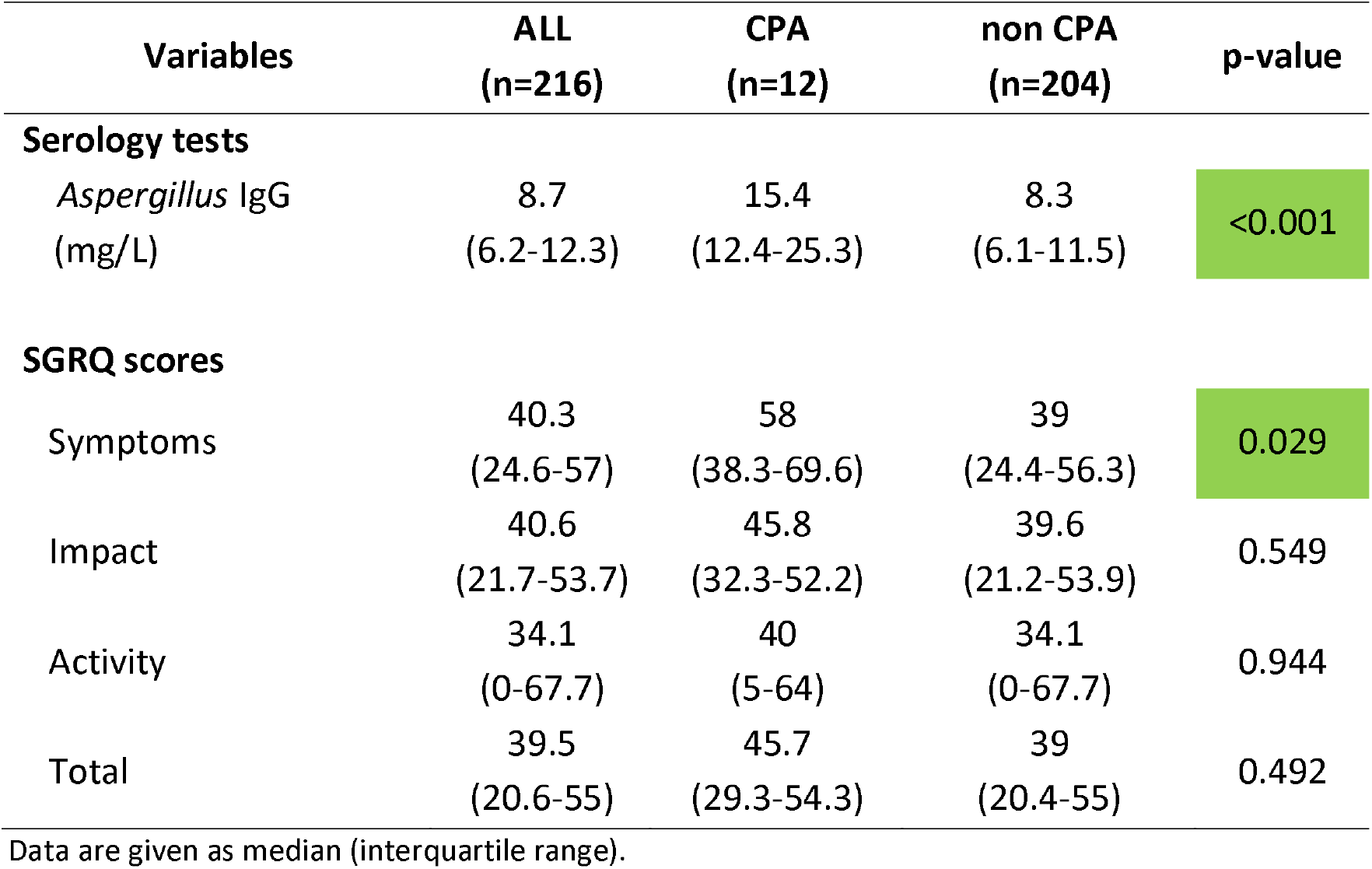
IgG tests and SGRQ results at baseline.

The overall prevalence of *Aspergillus*-specific IgG positivity was 30% (64/216) at baseline. All (n=12) patients from the CPA group had positive levels of *Aspergillus*-specific IgG. In the non-CPA group, 52 (26%) patients showed positive *Aspergillus*-specific IgG. There was a statistical significance between positive *Aspergillus-*specific IgG across CPA and non-CPA groups [p <0.001, median 15.4 (CPA) vs 8.3 (non-CPA)] (Table 2).

Nine (4%) patients died before their end of TB therapy appointment. Eight of them (90%) had extensive infiltrates and/or cavitation in both lungs. Immulite *Aspergillus*-specific IgG level was positive in four (44%) of these patients. Chronic kidney diseases (n=3, 33%) and diabetes mellitus (n=1, 11%) were linked to mortality. One possible CPA patient with multicavitary TB disease died with no other underlying disease. There were no deaths in proven and probable CPA patients.

At the end of TB therapy, those assessed (n=128) had a similar spectrum and frequency of symptoms with the exception of fatigue (60% vs 13%, p=0.001) and cough (50% vs 11%, p=0.005) which were more common in CPA (Table 3). The overall prevalence of *Aspergillus*-specific IgG positivity was 52% (66/128). Ten of 128 patients had CPA, an incidence of 7.8% (95%CI: 7.9, 20.4). All (n=10) patients from the CPA group had positive levels of *Aspergillus*-specific IgG. In the non-CPA group, 45 (42%) patients showed positive *Aspergillus*-specific IgG. Immulite *Aspergillus*-specific IgG level increased in CPA patients compared to non-CPA patients [p<0.001, median 24.2 (CPA) vs 9 (non-CPA)]. Quality of life scores dropped dramatically during TB therapy, reflecting improved well being, from a median of 35.9 to 4.8 for overall score (p<0.001) (Table 4). However, this drop was smaller in those with proven CPA (62% decrease in SGRQ total score) than in those without CPA (91% decrease in SGRQ total score). Radiological findings improved during TB therapy with the exception of pleural thickening, and the interval development of one aspergilloma (Table 3). There was also a slight increase in bronchiectasis (on plain radiography).

**Table 3.**
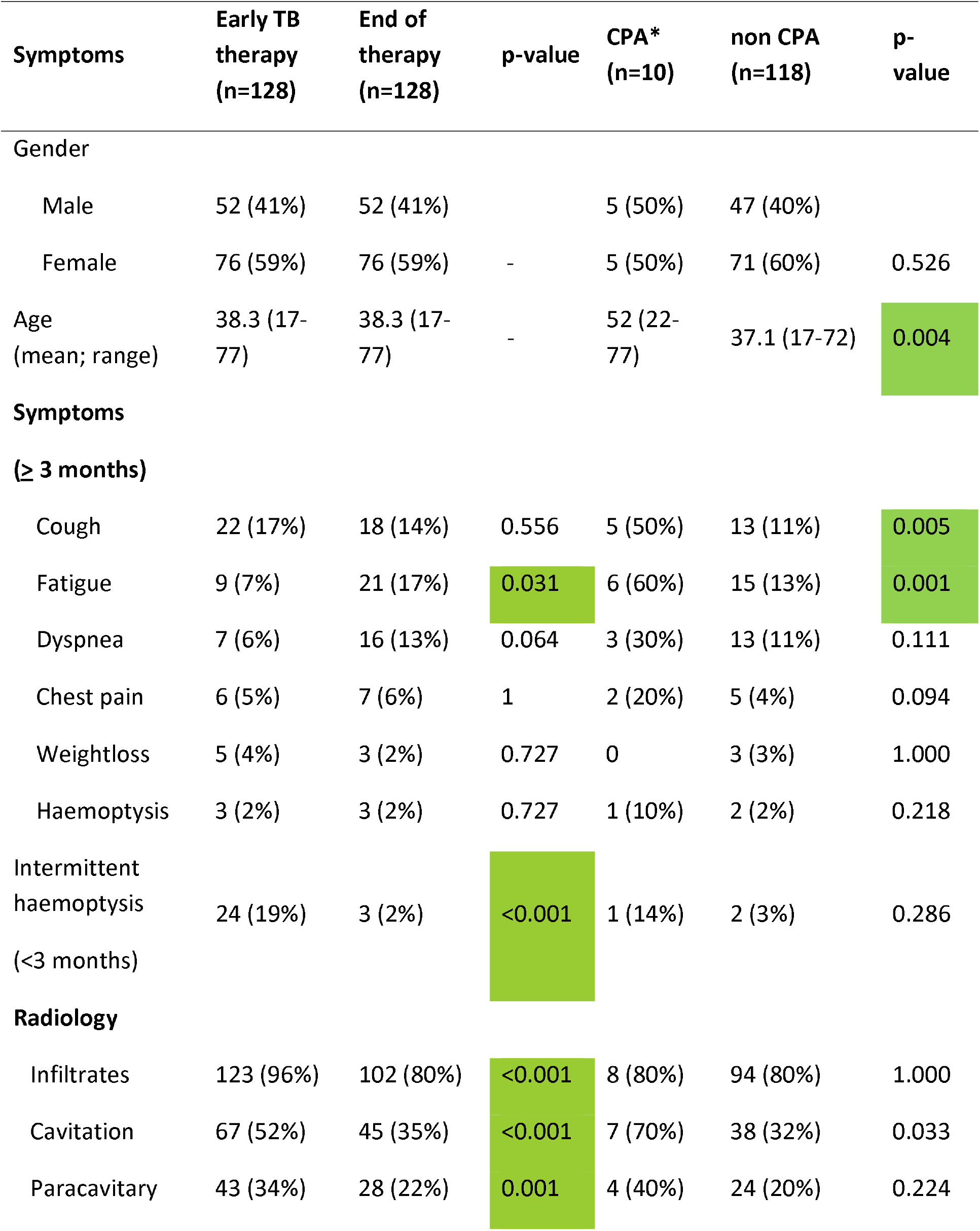
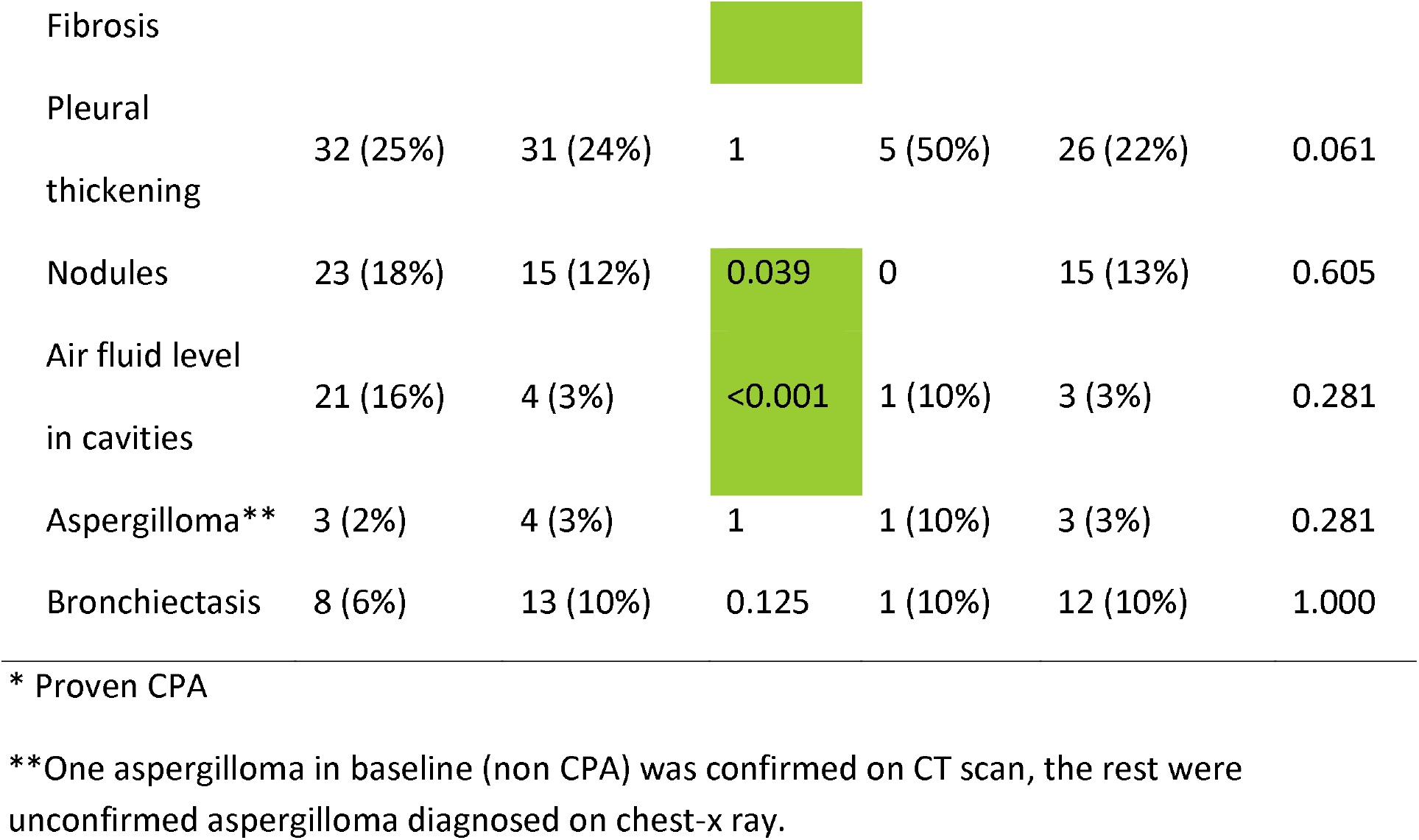
Results for end of TB therapy assessment compared to baseline for all those assessed at both time points, using the CPA categorization at the second time point.

**Table 4.** IgG tests and SGRQ comparisons for all patients assessed at baseline and at end of TB therapy, using the CPA categorization at the second time point.

| Variables | Early TB therapy<br>(n=128) | End of therapy<br>(n=128) | p-value | CPA<br>(n=10) | non CPA<br>(n=118) | p-value |
| --- | --- | --- | --- | --- | --- | --- |
| **Serology tests** |  |  |  |  |  |  |
| *Aspergillus*-specific IgG (mg/L) | 9.2<br>(6.5-12.7) | 9.7<br>(6.9-14.2) | 0.078 | 24.2<br>(13.8-76.1) | 9<br>(6.6-13.1) | <0.001 |
| **SGRQ scores** |  |  |  |  |  |  |
| Symptoms | 37.2<br>(18.4-56.7) | 3<br>(0-22.5) | <0.001 | 23.3<br>(9.6-37.3) | 3<br>(0-21.1) | 0.002 |
| Impact | 39<br>(21.5-51.9) | 4<br>(0-13.6) | <0.001 | 9.2<br>(5-37.7) | 3.7<br>(0-13.1) | 0.007 |
| Activity | 21.9<br>(0-59.4) | 0<br>(0-8.4) | <0.001 | 23.6<br>(0-53.9) | 0<br>(0-0) | 0.002 |
| Total | 35.9<br>(16.3-51.4) | 4.8<br>(4-12.8) | <0.001 | 17.4<br>(8-36.8) | 3.4<br>(0-12.2) | 0.003 |

Data are given as median (interquartile range).

Of the 12 patients with features of CPA at baseline, seven (58%) were categorised as non-CPA cases at the end of TB therapy without any antifungal therapy or surgery. This was mainly due to decrease in symptoms and in infiltrates seen in chest x-rays (Figure 2). Two (17%) were lost to follow-up and not assessed. The remaining three patients continued to have CPA features at the end of TB therapy. An additional seven patients developed CPA during their TB therapy, resulting in a total of ten (8%) CPA cases at the end of TB therapy. The risk for developing CPA during TB therapy appears to increase with age [p=0.004, mean 52 (CPA) vs 37.1 (non-CPA)] Table 3.

**Figure 2.**
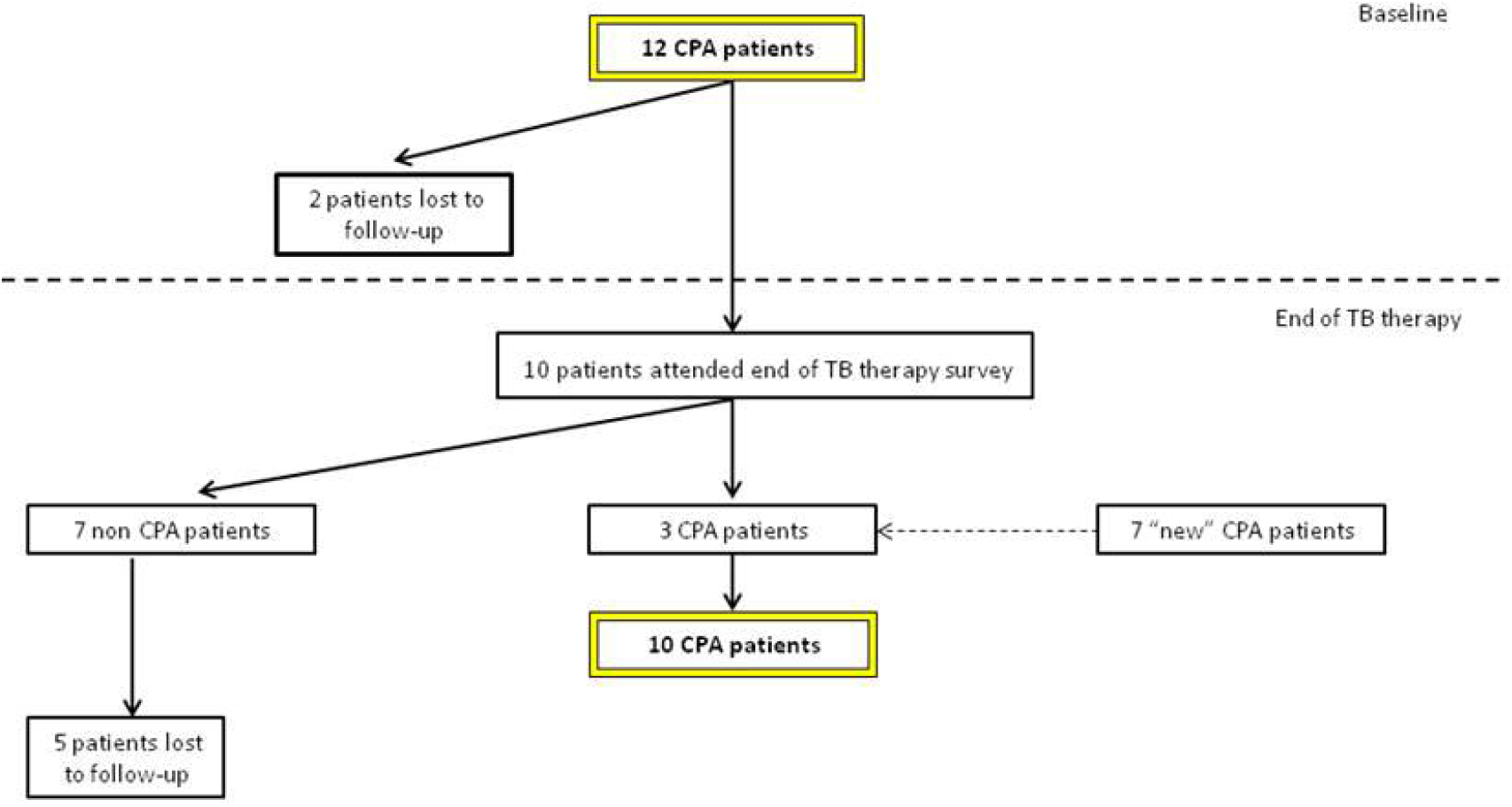
Chronic pulmonary aspergillosis patients during two time-point assessments

Of five probable CPA cases at baseline, two improved clinically (resolution of symptoms) and one patient became a possible CPA with decreased persistent symptoms but stable pleural thickening by the end TB therapy. Two patients were lost to follow-up. Of 15 possible CPA at the baseline, one patient improved clinically, another met the criteria for probable CPA and three patients did not have features of CPA cases at the end of TB therapy assessment; 10 patients were lost to follow-up. At the end of TB therapy, probable CPA and possible CPA were detected in seven (5%) patients and four (3%) patients, respectively. The clinical and radiology profile of patients at both time-points are compared in Table 3 and 4.

## Discussion

The incidence of proven and probable CPA was 7.9% (95%CI: 4.7, 12.3) at baseline to 13.3% (95%CI: 7.9, 20.4) at the end of TB therapy, although the latter figure is marred by substantial loss of follow up, including some deaths, probably of CPA, between these 2 time points. There are three confounding factors that need to be considered when interpreting this finding. First, different patients met the criteria for CPA diagnoses at baseline and end of TB therapy. Second, a notable number of patients were lost in follow-up. Third, the accuracy of the CPA diagnosis is a challenge with 5 probable and 15 possible CPA patients at baseline and 7 probable and 4 possible at the end of TB therapy. Furthermore, some patients had aspergillomas visible on their chest radiograph without clinical or serological features of CPA; 5 at baseline and 1 at the end of TB therapy. If all these patients are assumed to have CPA (which is unlikely for the possible group), the incidence of CPA was 17% both at baseline and at the end of TB therapy. Nevertheless, it is likely that the true prevalence of CPA during the course of treatment of TB is somewhere between 6% and 17% at baseline and 8% and 17% at the end of therapy, in Indonesia. These are the first estimates of CPA incidence during anti-TB therapy, made globally.

One of the study objectives was to determine how many patients have dual infections. Co-infection of active pulmonary TB and CPA is reported in previous studies [6,27–29]. Our data indicates more frequent dual infection than previously thought. However, many of these patients appear to resolve their *Aspergillus* infection and had ‘self-resolving CPA’. Several other patients failed to attend the second assessment, so their fate is uncertain. The cause of death in those lost to follow up in our study was uncertain, including determining whether any dual infection patients died before assessment. In Japan, the term chronic progressive pulmonary aspergillosis was coined to separate those who deteriorated from those who remained clinically and radiological stable [21]. We have clear cut evidence of this divergence of disease behaviour in this study.

Male sex was an independent risk factor for CPA in this study. Earlier studies also showed male predominance in CPA compared to *Aspergillus* colonisation in Japan [30], and in a large CPA cohort in Manchester [25]. It can be speculated that this is due to higher prevalence of other risk factors such as smoking, but supportive data are lacking.

Prior studies have demonstrated the rate of CPA in post-TB therapy populations varies between 4.9%-14.3% [2,10]. In Nigeria the CPA rate was reported to be 8.7% at the end of TB-therapy [5] and in Uganda, 9% of HIV-infected patients had Aspergillus-specific IgG detectable at the end of TB-therapy [31]. A cross sectional study from Iran reported 13.7% of 124 HIV-negative TB patients developed CPA (76% patients were on active TB treatment) [4]. All of these CPA rates are within the ranges of our study. However, these studies provide no or minimal information about the status of the patients at baseline and reflect pooled patient populations assessed at varied time points after TB therapy [2,10].

One element of the diagnosis of CPA is symptoms for ≥3 months (weight loss and/or cough and/or haemoptysis) [15]. But as can be seen, this definition is challenged by our data. TB gives rise to weight loss and cough and many patients have some degree of haemoptysis. We found the relative frequency for CPA and TB to be: cough 93% vs 13%, fatigue 42% vs 8% and weight-loss 17% vs 1% respectively (Table 1). Three months of cough is more discriminatory than any cough. Fatigue has been studied and a score devised for COPD patients applied to CPA patients appears to be useful, but has yet to be translated into routine clinical practice [32]. Chest pain appeared to be a useful discriminator in Nigeria but was only present in 25% of CPA patients [5]. Three months of haemoptysis was found in only 8% of patients with CPA, compared to 1% with TB, but shorter periods of haemoptysis were common and not very discriminatory (Table 1).

In addition to specific symptoms, the SGRQ quality of life score was recorded at baseline. Unremarkably for patients with tuberculosis, scores were moderately high (Table 2), but not at all discriminatory between those with only TB and those with CPA and TB, at baseline. However at 4-6 months after starting anti-tuberculous therapy, those with CPA had persistently raised SGRQ scores in all domains, and there was almost complete resolution of all symptoms in those without (Table 4). Therefore this score might be very useful in Indonesia for identifying CPA at the end of anti-TB therapy. It is worth noting that the SGRQ scores in this study are remarkably low compared with other patients who have recovered from TB in the USA [33], but not Papua, Indonesia [34].

The radiological appearances of CPA and TB partially overlap, with upper lobe predominance and cavitation. While TB does not usually cause pleural thickening in the apices, consolidation can occur which can appear like pleural thickening. Likewise peri-cavitary infiltrates are very typical of CPA, but local consolidation can mimic these findings too. The first description of an aspergilloma by Deve in 1938 described a huge cavity with a fungal ball, which is un-mistakable, but more subtle findings are now understood to be typical of CPA [35]. The typical appearance of a fungal ball or aspergilloma includes the air crescent or air meniscus sign, first described in the context of pulmonary hydatid disease [36,37]. Our study lacks confirmation from CT of the findings seen on chest radiographs in 97.7% of patients. This is a clear cut study weakness and arises primarily from a lack of funds to afford CT scanning for most patients and an inability to identify in real time those whose disease required better anatomical definition with CT, as serology was done in batches months after the patients were seen.

There is intrinsic uncertainty in the interpretation of chest radiographs. Concordance between readers is far from 100%, both in ascertaining cavitation in TB (50-70%) and in aspergillosis [38,39]. Our radiographs were read by an experienced radiologist in Indonesia (where the only aspergillosis radiological diagnosis made currently is an aspergilloma) with confirmation by a highly experienced clinician focussed on pulmonary aspergillosis. Separating CPA from TB with a chest radiograph alone with any certainty is not possible.

Partly using this dataset (end of TB therapy only), and a separate cross-sectional study population (n=71) and both normal and disease controls (n=190), we derived the optimum cut-off for *Aspergillus*-specific IgG (11.5mg/L) in Indonesia for the Immulite assay [26]. The antibody titres varied substantially between both time points, indicative of a complex interaction between *Aspergillus* and the immune system during the months of anti-TB therapy.

The presence of an aspergilloma is highly indicative of CPA, with echinococcus and necrotizing lung tumours really the only differential diagnoses. An aspergilloma may represent a single aspergilloma, without symptoms or be a feature of chronic cavitary pulmonary aspergillosis. In this study, aspergillomas were detected in 11 patients, with 8 (72.7%) patients having positive PCR TB and/or AFB smears at baseline, indicating co-infection of *Aspergillus* in active TB patients. Five (45.5%) patients had positive *Aspergillus*-specific IgG by Immulite, four (36.4%) patients in the end and/or post TB therapy and one (9.1%) patient showed persistent positive *Aspergillus-*specific IgG by Immulite but with no accompanying symptoms. Two patients had CT scan-confirmed aspergilloma, both with bacteriogically confirmed TB.

In most patients with aspergillomas, *Aspergillus* IgG was negative when first seen and most were therefore classified as non-CPA. Seroconversion of Immulite from negative to positive *Aspergillus-*specific IgG occurred in all 5 patients with persistent aspergilloma. Aspergillomas with negative *Aspergillus fumigatus* IgG antibody has been observed in Africa and may relate to non-fumigatus infections or a muted antibody response [2,5].

The other study objective was to determine how many patients are mis-diagnosed as TB when in fact they have CPA. Ruling out TB can be difficult, so complete confidence in this assertion for individual patients is usually difficult. Community-acquired *Aspergillus* community acquired pneumonia without any evidence of immunocompromised can occur [40,41], often following significant environmental exposures such as bark chippings, compost and gardening [42,43]. Some of these patients fail to complete resolve their *Aspergillus* infection, and do not die developing CPA. It is highly likely that some of the clinically diagnosed cases of TB in this study with positive *Aspergillus* IgG had community-acquired *Aspergillus* CAP and some went on to develop CPA.

We therefore propose that CPA should be included as an alternative diagnosis of pulmonary TB, especially if there is no clinical and radiological improvement after starting TB therapy. Pulmonary TB and CPA may co-exist and both diseases require their own management plan. In the context of non-tuberculous mycobacterial infection and CPA, the outcomes of those with dual infections were much worse (11.5-fold increased mortality) [19], this has yet to be ascertained for those with TB. The availability of *Aspergillus* antibody testing and CT scanning are important components of optimal CPA diagnosis. The study is important for the future study of CPA in Indonesia. CPA as an alternative or co-existing disease in TB patients in Indonesia is important for clinicians and to galvanise social and political will of multiple stakeholders to support improvement of diagnostic capacity of fungal disease in Indonesia.

## Data Availability

All data are archived on University of Manchester servers are are available on request

## Acknowledgement

We would like to thank all the staffs at the Department of Parasitology Faculty of Medicine University of Indonesia for their help in sample storage and testing of sputum samples. We gratefully acknowledge Dr. John Belcher of the department of medical statistics for statistical support. We would also like to acknowledge support from the staffs at the Mycology Reference Centre Manchester for providing training of manual *Aspergillus-*specific IgG ELISA tests and support throughout the project.

## Contributors

FS, AR, EB and RW contributed to the conception and design, acquisition of the data, and analyses and interpretation of the data. RA, RS, MT, CYIS, FN, JR, ARA, DH, and MCR contributed to acquisition of the data, and analyses and interpretation of the data. RR and DWD contributed to the conception and design, analyses and interpretation of the data. All authors made a significant intellectual contribution to the drafting of the work and contributed to the approval of the final version of the manuscript.

